# COVID-19 first and delta waves in relation to ACEI, ARB, Influenza vaccination, and comorbidity in a North Metropolitan Barcelona Health Consortium

**DOI:** 10.1101/2021.11.17.21265440

**Authors:** Marta Juanes-González, Ana Calderón-Valdiviezo, Helena Losa-Puig, Roger Valls-Foix, Marta González-Salvador, Marc León-Pérez, Luís Pueyo-Antón, Celia Lozano-Paz, Maite Franco-Romero, Josep Vidal-Alaball, Anna Puigdellívol-Sánchez

## Abstract

**BACKGROUND:** Some authors have reported that angiotensin converter enzyme inhibitors (ACEI) and angiotensin receptor blockers (ARB) improve clinical outcomes in hypertensive COVID-19 patients, and others have proposed cross-protection for influenza vaccination. This study explores the impact of these variables on the evolution of hospitalized patients, focusing in the first wave and the Delta wave.

**METHODS:** Hospitalizations (n=1888) from March 1, 2020, to July 31, 2021, in the Hospital of Terrassa, the referral center for the free access Terrassa Health Consortium in the North Metropolitan Barcelona Health Region (population=167,386) were studied. The number of chronic treatments and conditions of patients from the initial outbreak (n=184) and the Delta outbreak (n=158) were recorded.

**RESULTS:** Of the non-survivors, 96.3% were aged >60 years in the first wave and 100% were aged >70 years in the Delta wave. In non-survival hospitalized patients aged >60 years, the percentage treated with ACEI was similar to general population but was significantly different for ARB treatments of influenza vaccination, although associated to a higher comorbidity and age. In July 2021, the number of hospitalizations for patients aged <50 years was higher than March 2020 and 22% of hospitalized patients without chronic treatments and conditions needed admission to the intensive care unit. Mortality was reduced in the groups with most comorbidities who received influenza and SARS-CoV2 vaccination.

**CONCLUSIONS:** In COVID-19 infection, age and comorbidity are related to survival, ACEI use is safe. A high proportion of patients without comorbidity require hospitalization and intensive care.

## INTRODUCTION

Mortality rates in Barcelona (Catalonia, Spain, EU) suddenly tripled on March 15, 2020 [1], two weeks after the COVID-19 outbreak in Italy [2] and four months after the COVID-19 outbreak in China. This made it the third country worldwide to have a mass outbreak in the first wave. After several waves along 2020 and 2021, cases again began to rise suddenly at the end of June 2021, this time in relation to the Delta variant.

Potential interventions to treat the disease had been proposed early in the pandemic [3]. Angiotensin-converting enzyme inhibitor (ACEI) and angiotensin receptor blocker (ARB) use was reported to improve clinical outcomes in patients with COVID-19 and hypertension [4], but this was only supported by sparse experimental data showing the protective effect of an ACEIs on neurogenic pulmonary edema [5]. Some concerns were raised about this use given that these medications may raise expression of ACE2, the receptor for severe acute respiratory syndrome-coronavirus 2 (SARS-CoV-2); however, these concerns later proved to be unfounded [6.7]. The use of influenza vaccine was also suggested [3], with some authors hypothesizing the potential for cross-protection [8] and others reporting primary evidence in support of this proposal [9]. Influenza virus and coronavirus use similar strategies through hemagglutinin esterases to engage sialoglycans at the surface of target cells [10,11].

This study aimed to assess the influence of these different factors—influenza vaccination, ACEI therapy, and ARB therapy—on the evolution of SARS-COVID-19 infection. The prevalence of those factors was compared between hospitalized patients and the general population. Detailed data are also presented from the first and the Delta waves in relation to those factors, associated comorbidity, and treatments.

## METHODS

The study was approved by the ethics committee of the Terrassa Health Consortium (THC) on April 8 (ref 02-20-161-021) and the observational clinical trial was posted on April 29 (NCT 04367883). The planning, conduct, and reporting of the study were in line with the principles of the Declaration of Helsinki.

The THC is a free public integral health organization in the North Metropolitan Barcelona Health Region. This region includes seven primary healthcare centers, one long-term care center, and the Hospital of Terrassa, which serves as a referral center for hospitalization (n = 167,386; aged >60, n = 34,928).

### Quantification of hospitalizations

The Data Analysis Control Department collected anonymized data for COVID-19 cases and hospitalized patients since March 1 2020, together with data about prior treatment with ACEI and ARB, prior influenza vaccination, and survival. The presence of COVID-19 was determined either by polymerase chain reaction (PCR), antigen test, or clinical criteria (compatible interstitial pneumonia) from March 1, 2020, to July 31, 2021.

### Comparison between hospitalized patients and the general population

In the first wave (March 1 to June 30, 2020), 555 hospitalized patients with COVID-19 were compared to those in the general population with regard to the percentages with influenza vaccination, ACEI therapy, and ARB therapy. This process was repeated for the Delta wave.

The Chiromas® (Seqirus S.r.l). and Chiroflu ® (Seqirus S.r.l) vaccines, which have similar compositions, were used to vaccinate adults against seasonal influenza in 2019–2020 and 2020–2021. Vaxigrip Tetra (Sanofi Pasteur Europe) was also used in the 2020–2021 season. COVID-19 vaccinations with the Pfizer^®^, Astra-Zeneca^®^, Moderna^®^, and Janssen^®^ vaccines began in 2021, starting with older people. Vaccination is considered complete if one dose of Janssen^®^ or two doses of another vaccine has been received.

### Comorbidity

The number of chronic treatments, conditions, and admissions to the intensive care unit (ICU) for PCR-positive patients hospitalized in the THC zone (n = 184) during the first wave of COVID-19 when mortality began to rise (March 10 to 25, 2020) were recorded. The variables were again recorded for patients hospitalized in the THC zone (n = 158), when hospitalization suddenly increased again due to the Delta variant and a night curfew was imposed (July 16 to August 20, 2021). These data were obtained from medical records. The types of chronic treatments were classified according to the Anatomical Therapeutic Chemical (ATC) classification system [12].

### Statistical analysis

Chi-Square tests were used to compare the percentage of treatments between hospitalized patients and the general population aged >60 years. Independent student *t*-tests were used to compare absolute values between independent groups.

## RESULTS

### Quantification of the hospitalizations

The distribution of patients hospitalized with COVID-19 (n = 1881 from the THC zone), together with the twice monthly mortality, is shown in Figure 1 for the period from March 1, 2020, to July 31 2021. Overall mortality in hospitalized patients fluctuated during this period from 18.1% during the first wave (March–June, 2020), to 8.5% in the second wave (October–November, 2020), to 15.2% between December 2020 and February 2021, and to 4.3% after March 2021. However, mortality remained low in July, at 4.4%, despite the sudden increase in hospitalizations.

**Figure 1.**
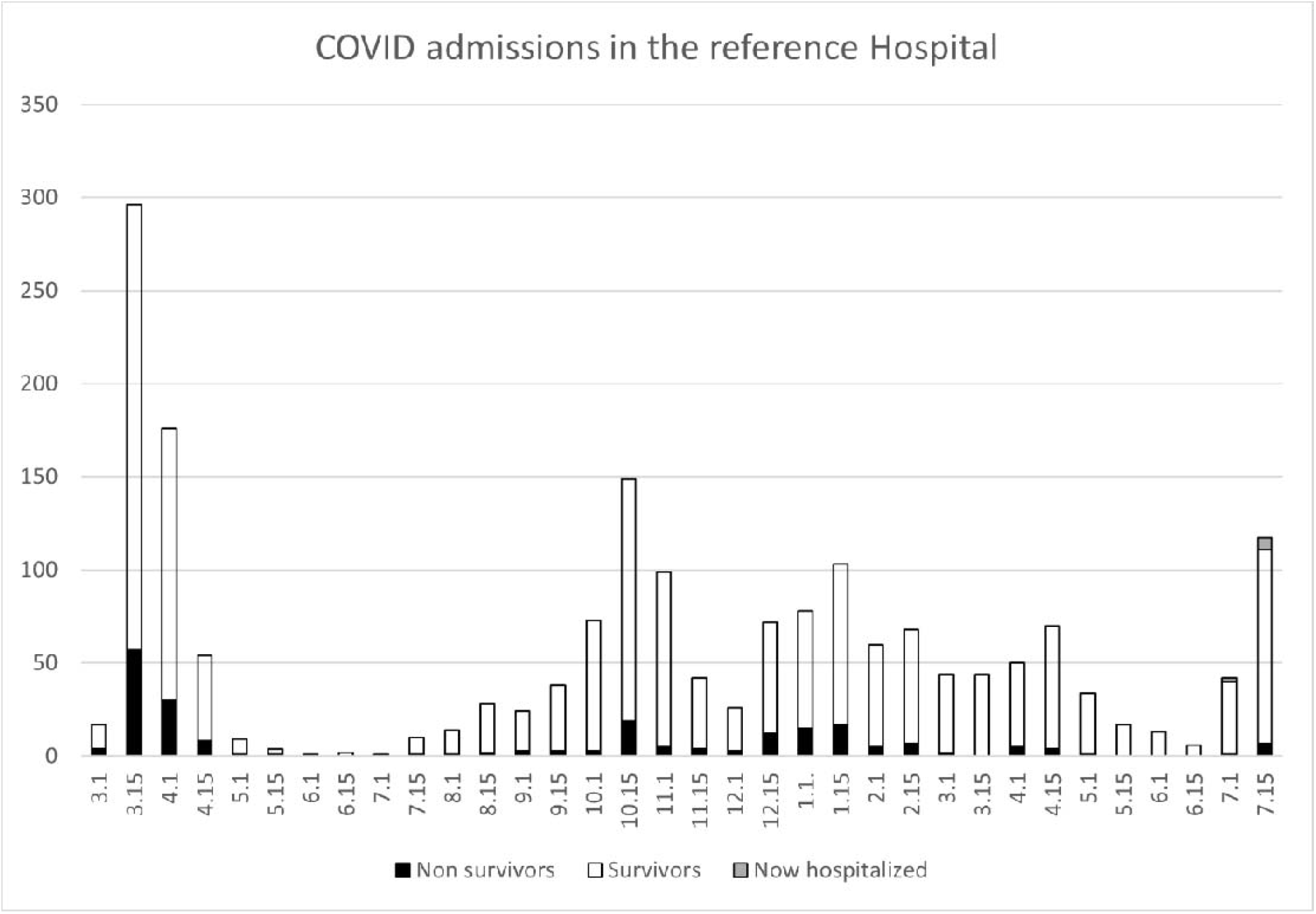
Hospitalized COVID-19 cases in the THC zone at half-month intervals from March 1, 2020, (3.1) to July 31, 2021.

Hospitalization and survival rates in the first month of the COVID-19 outbreak (March 2020) and the first month of the Delta outbreak (July 2021) are compared in Figure 2.

**Figure 2.**
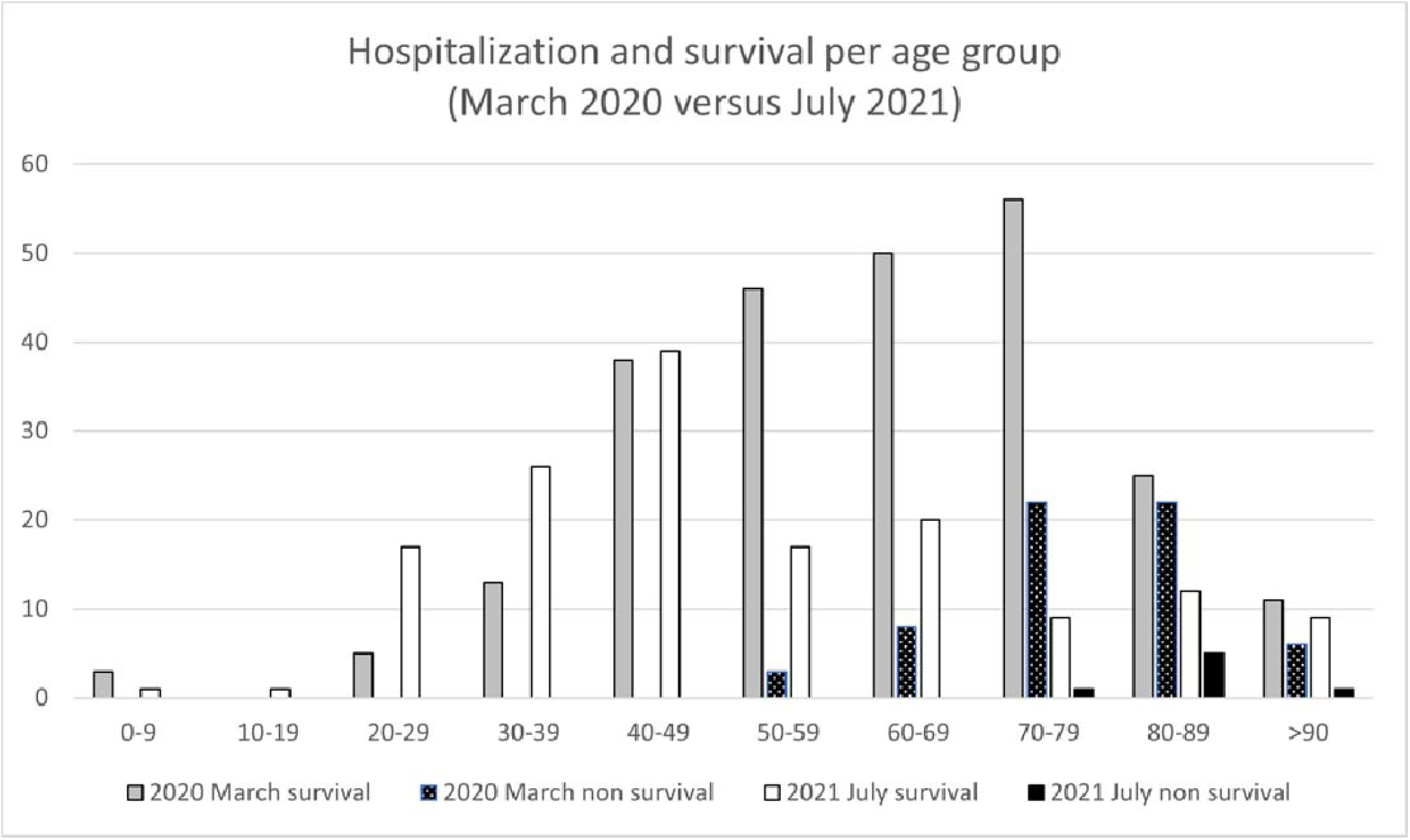
Incidence of hospitalizations and survival per age groups in the first full month of the first wave (March 2020) and of the Delta wave (July 2021).

Most non-survivors (96%) were aged >60 years in the first wave (Table 1), and all were aged >70 years in the Delta wave (Figure 2).

**Table 1.** COVID-19 hospitalizations for the THC zone in the first wave by age, gender, ACEI, ARB, and Influenza vaccination in comparison with the reference population aged >60. Detailed *p* is shown for significant differences only (*).

|  |  | Age | Gender<br>% Males | ACEI | ARB | Influenza<br>vaccination |
| --- | --- | --- | --- | --- | --- | --- |
| Hospitalized COVID-19 cases<br>(March 1 to June 30) |  |  |  |  |  |  |
| Overall<br>(n = 555) | Non-survivors (n = 108) | 80.5 ± 10.7 | 58.3% (63) | 28.7% (31) | 20.4%<br>(22) | 62% (67) |
|  | Survivors (n = 447) | 63.5 ± 17.4 | 51.7% (364) | 23.3% (104) | 9.4%<br>(42) | 35.6% (159) |
| Reference population (n = 167,386) |  |  |  | (n = 16,242) | (n = 1812) | (n = 21,456) |
| Aged >60<br>(n = 363,<br>65.4%) | Non-survivors<br>(n = 104, 96.3%) | 81.5 ± 9.5 | 56.7% (59) | 27.9% (29) | 21.2%*<br>(*p* = 0.006)<br>(22) | 64.4%*<br>(*p* = 0.0001)<br>(67) |
|  | Survivors<br>(n = 259, 57.8%) | 75.6 ± 9.7 | 48.3% (126) | 30.1% (78) | 13.9% (36) | 51.4%*<br>(*p* = 0.044)<br>(133) |
| Reference population aged >60 (n = 34,928) |  |  |  | 34.7%<br>(n = 11,501) | 11.5%<br>(n = 3845) | 44.9%<br>(n = 15,703) |

More hospitalizations were required for patients aged <50 years old in July 2021 compared with March 2020 (Figure 2), including many patients who were not taking a chronic drug (N = 59) (Table 2).

**Table 2.** Comorbidity and survival. Comorbidity (number of chronic treatments and pathologies), age, and gender, related to ACEI and ARB treatments, influenza vaccination, or SARS-CoV-2 vaccination in 2021, length of hospital stay, admission in Intensive Care Unit (ICU), and survival in the initial COVID-19 PCR+ hospitalized cases (March 10-25 admissions) and during the Delta wave peak (July 2021).

| Chronic treatments | Chronic conditions | Age | Gender, % females | % ACEI | % ARB | Influenza vaccination | SARS-CoV-2 vaccination (complete) | Length of hospital stay | ICU | Non-survivors |
| --- | --- | --- | --- | --- | --- | --- | --- | --- | --- | --- |
| >8 (n = 28) | 11.6 ± 5.4 | 77.9 ± 19.5 | 60.7% | 25.0% | 14.3% | 85.7% | 96.4% (92.8%) | 12.0 ± 7.5 | 0.0% | 7.7% |
| 5–7 (n = 18) | 7.1 ± 3.5 | 70.0 ± 16.2 | 55.6% | 33.3% | 29.4% | 55.6% | 44.4% (27.8%) | 8.4 ± 4.7 | 22.2% | 19.0% |
| 2–4 (n = 33) | 5.1 ± 2.6 | 54.0 ± 17.8 | 48.5% | 12.1% | 12.1% | 30.3% | 42.4% (33.3%) | 9.5 ± 5.0 | 9.1% | 3.0% |
| 1 (n = 20) | 3.4 ± 2.6 | 44.3 ± 15.9 | 45.0% | 10.0% | 5.0% | 5.0% | 25.0% (10.0%) | 5.1 ± 3.0 | 10.0% | 0.0% |
| 0 (n = 28) | 2.6 ± 2.2 | 42.1 ± 12.3 | 35.7% | 3.6% | 0.0% | 10.7% | 35.7% (7.1%) | 9.9 ± 6.4 | 28.6% | 3.6% |
| 0 (n = 31) | 0.0 | 33.8 ± 7.3 | 38.7% | 3.2% | 0.0% | 3.6% | 26.7% (3.3%) | 6.6 ± 5.4 | 22.6% | 0.0% |
| **March 10 to 22 2020** |  |  |  |  |  |  | **Charlson index** |  |  |  |
| >8 (n = 49) | 4.9 ± 2.2 | 75.2 ± 11.6 | 49.0% | 20.4% | 24.5% | 55.1% | 3.0 ± 2.7 | 16.0 ± 16.0 | 10.2% | 57.1% |
| 5–7 (n = 42) | 3.4 ± 1.7 | 70.6 ± 10.4 | 35.7% | 31.0% | 21.4% | 54.9% | 1.3 ± 1.5 | 17.6 ± 15.7 | 22.0% | 31.0% |
| 2–4 (n = 53) | 2.4 ± 1.3 | 63.6 ± 15.2 | 43.4% | 38.5% | 0.0% | 30.2% | 0.6 ± 1.1 | 18.3 ± 20.0 | 23.5% | 17.0% |
| 1 (n = 13) | 2.2 ± 1.5 | 63.6 ± 11.8 | 23.1% | 15.0% | 0.0% | 38.5% | 0.5 ± 0.7 | 12.4 ± 10.2 | 17.0% | 23.0% |
| 0 (n = 11) | 1.4 ± 0.5 | 52.2 ± 19.8 | 36.4% | 0.0% | 9.1% | 27.3% | 0.2 ± 0.6 | 15.2 ± 13.1 | 36.0% | 18.0% |
| 0 (n = 16) | 0.0 | 42.5 ± 15.2 | 31.3% | 0.0% | 0.0% | 12.5% | 0.0 ± 0.0 | 12.9 ± 16.1 | 5.0% | 0.0% |

### Comparison between hospitalized patients and the general population

Among people aged >60 years in the first wave, no significant differences were found in ACEI usage between hospitalized PCR-positive COVID-19 cases and in the general population. However, there was a significant difference in ARB use and influenza vaccination (p<0.05, Table 1), with a close association with more comorbidity and older age (Table 2).

During the Delta wave, influenza vaccination rates varied from 30% to 85.7%, but the mortality remained below 10% in most groups with different comorbidities. The highest ARB use was associated with patients who had a high number of chronic conditions but a lower percentage of complete SARS-CoV2 vaccination, including two non-vaccinated patients who died aged >80 years. Finally, ACEI use in all COVID-19 groups was less than that of the general population (Tables 2).

### Comorbidity

Among patients in the first wave with one pathology, 30% had hypertension; 21.4% had obesity, thyroid, or mental disorders; 7% had neurological disorders, osteoporosis, gastric, or colon pathologies; and 3.3% had chronic obstructive pulmonary diseases. Patients admitted in July 2021 with one pathology and no chronic treatments had backache (30.8%), obesity or dyslipidemia (15.4%), or other conditions (7.7%; these included sleep apnea, varicose veins, dyspepsia, Hodgkin lymphoma, or tinnitus).

Up to 91 different drugs were taken by patients admitted in the first wave. However, among patients with one pathology, 16.7% received analgesics (ATC code M01A), benzodiazepine treatments (N05B) or proton inhibitors (A02BC); 13.3% received diuretics (C03) or ACEI (C09); 6.7% received serotonin reuptake inhibitors (N06AB), calcium (A12A), betablockers (C07), other antiarrhythmics (C01B), direct thrombin inhibitors (B01AE), or ophthalmic prostaglandin analogs (S01EE); and 3% received another 13 different chronic drugs. In July 2021, patients using one treatment received the same agents (M01A, N05B, A02BC, C09, N06AB, C07, S01EE) and 5% used other 9 different types of chronic drugs.

During the Delta outbreak, all cases showing bilateral interstitial pneumonia also had a positive confirmatory antigen or PCR test. A non-significant trend was observed for a higher proportion of patients without comorbidity to require intensive care admission (22.6% vs 5%) even when those cases were younger (33.8 vs 42.5 years); more patients in comorbid groups were female in July 2021 (Table 3) but males were predominant in both waves (59.8% and 53.2% in March 2020 and June 2021, respectively).

## DISCUSSION

The official Spanish “state of emergency” began on March 14 when mortality rates began to increase markedly nationwide, reaching the mortality peak on March 25 [13]. The sample of patients around this initial lockdown period (March 10 to 25, 2020) is, therefore, a good indicator of the potential virulence of COVID-19 in the absence of countermeasures.

Overall mortality among hospitalized patients was 18.1% during the first wave (March– June, 2020), related to the B3a and B9 strains [14]. This fell to 8.5% during the second wave (October–November, 2020). It is difficult to discern if this was due to a new variant or an improvement in detection and early treatment. The 20E (EU1) variant was reported to originate in Spain as of summer 2020, before spreading to the rest of Europe [15]. The increase in mortality observed between December 2020 and February 2021 was probably related to the arrival of the alpha variant, initially detected in UK (also known as B.1.1.7) [16], which showed a quicker spread. Mortality then decreased to 4.3% after March 2021, probably due to the massive SARS-CoV-2 vaccination program in the older population. Overall mortality remained low in July, at just 4.0%, despite the arrival of the delta variant, which accounted for more than 70% of cases that month [17].

The rapid spread in Catalonia during July 2021 was probably enhanced by tourism, the crowded Saint John’s Day festivity on June 23, and the end of restrictions by June 26 (e.g., face masks, social meetings of <10 people, and geographical movement). These changes preceded the summer spread of the Delta variant around Europe and America.

### Hospitalizations

Only PCR-positive cases of interstitial pneumonia were considered in this study for March 2020, but there was other 17 hospitalized pneumonia in the same studied period that have not been considered here for being PCR-negative. However, all hospitalized cases in July with bilateral interstitial pneumonia had COVID-19 confirmed by antigen test or PCR, suggesting a higher degree of detection when both techniques were used in combination. There is preliminary evidence of false negative results associated with the antigen test alone [18,19]. The Alpha variant in the UK, which had already showed improved spread [16, 20], lead to suspected cases being pursued by the addition of further PCR if symptoms persisted, according to current protocols.

The male predominance in hospital admission is consistent with that reported elsewhere [21]. All hospitalized patients, except those with multiple comorbidities (>8 chronic treatments), had vaccination rates below 50% during the Delta outbreak, which is less than the vaccination rate in the general population at the beginning of the summer [22]. Among patients taking >8 chronic treatments, who were aged 77.9 ± 19.5 years on average, 96% had completed their vaccination course. Of note, mortality in that group was reduced to 7.7% in the delta wave from 57% in the first outbreak, suggesting the protective role of the vaccination in this frail population.

### Relationship of ACEI and ARB use and of influenza vaccination

The comparable percentages of ACEI treatments in PCR-positive cases indicates that this treatment does not predispose patients to COVID-19 [6, 7]. finding is consistent with previous publications on improved COVID-19 outcomes in patients with hypertension [4] and with others regarding any association with hospitalization or death [23]. Although the percentages using ARBs or receiving the influenza vaccination were higher in this series, a clear relationship with increased comorbidity and age was evident. It is these latter factors that are the most likely reasons for death.

If influenza vaccination had shown clear cross-protection with COVID-19, we would expect a lower percentage of influenza vaccination in hospitalized cases than in the general population. Although the percentage of vaccinated patients was higher in these cases, this likely reflects a selection bias for vaccination in patients with more comorbidities [24]. Other groups, have recently described a protective effect of influenza vaccination in patients with SARS-CoV-2 infection [25]. Crossed immunization for COVID-19 by the influenza vaccine has not been proved in this study, but the reduced mortality in hospitalized patients with most comorbidities in July 2021 at least suggests the safety of combining the vaccines given that most patients will have received an influenza vaccine, a SARS-CoV-2 vaccine, or both. Furthermore, the role of the influenza vaccine in preventing co-infections is still of interest given the impossibility of predicting influenza behavior this coming winter after its low incidence in the 2020–2021 season [26].

Hospitalizations since the first wave have been influenced by preventive efforts to maintain a safe distance, wear masks, and limit visits in residential populations. SARS-CoV-2 vaccination has also clearly affected hospitalizations in older people during 2021. The mass influenza vaccination program for the elderly in the 2020–2021 season, combined with the low circulation of influenza viruses that winter, precludes conclusions about the role of potential cross-protection by the influenza vaccination.

COVID-19 vaccination rates in Spain among people aged >20 years averaged >90% at the end of August, with 75% of the Spanish population having received at least one dose and 65% being fully vaccinated. However, these have increased from the beginning of July, when these levels were 55.3% and 39.3%, respectively [22]. These data help to explain the overall reduction in hospitalizations.

### Comorbidity in respiratory patients

There was a trend to need hospitalization for COVID-19 disease among patients with hypertension and obesity. However, the great variability in chronic conditions and treatments, and the need for hospitalization among many patients with no prior pathology, suggest that severe disease may appear without comorbidity. Our results are consistent with other data in Europe comparing the Charlson comorbidity index in patients with and without COVID-19 [27] or influenza. Less comorbidity has been reported in patients with COVID-19 [28], but this may be influenced by the heterogeneity of the coronavirus strains circulating in a given area [29].

### Limitations

One limitation of the study is the lack of a gold standard technique for confirming COVID-19. False-negative rates of 38% on the day of symptom onset and 20% three days later have been reported [30] during 2020. Consequently, other than when analyzing the relationship with vaccination and comorbidity, we only included PCR-positive cases (Table 2). In 2021, all cases had positive antigen or PCR tests, suggesting that the diagnostic techniques have improved this year in real-world setting. Another limitation is that a second private hospital services the same health region as the THC, meaning that hospitalization rates could be underestimated. Nevertheless, it is feasible that the patients diagnosed in the other facility were comparable to our own.

## CONCLUSIONS

We are aware of no other study describing hospitalization rates for COVID-19 receiving ACEI, ARB, or prior influenza vaccination related to other chronic conditions and treatments comparing the First and Delta waves. Each of these variables was safe, and although cross-immunization for COVID-19 by the influenza vaccine was not proven, there is still interest in its role preventing co-infection. Moreover, the SARS-CoV-2 delta variant may lead to intensive care admission in more than 20% of hospitalized patients with no prior comorbidity.

## Data Availability

All data produced in the present study are available upon reasonable request to the authors

## ACKNOWLEDGMENTS

Lluís Valls-López and Michael Maudsley collaborated in the final revision of English spelling.

